# Social Capital Dimensions are Differentially Associated with COVID-19 Vaccinations, Masks, and Physical Distancing *

**DOI:** 10.1101/2021.09.13.21263543

**Authors:** Ibtihal Ferwana, Lav R. Varshney

## Abstract

**Background:** Social capital has been associated with health outcomes in communities and can explain variations in different geographic localities. Social capital has also been associated with behaviors that promote better health and reduce the impacts of diseases. During the COVID-19 pandemic, social distancing, face masking, and vaccination have all been essential in controlling contagion. These behaviors have not been uniformly adopted by communities in the United States. Using different facets of social capital to explain the differences in public behaviors among communities during pandemics is lacking.

**Objective:** This study examines the relationship among public health behavior—vaccination, face masking, and physical distancing—during COVID-19 pandemic and social capital indices in counties in the United States.

**Methods:** We used publicly available vaccination data as of June 2021, face masking data in July 2020, and mobility data from mobile phones movements from the end of March 2020. Then, correlation analysis was conducted with county-level social capital index and its subindices (family unity, community health, institutional health, and collective efficacy) that were obtained from the Social Capital Project by the United States Senate.

**Results:** We found the social capital index and its subindices differentially correlate with different public health behaviors. Vaccination is associated with institutional health: positively with fully vaccinated population and negatively with vaccination hesitancy. Also, wearing masks negatively associates with community health, whereases reduced mobility associates with better community health. Further, residential mobility positively associates with family unity. By comparing correlation coefficients, we find that social capital and its subindices have largest effect sizes on vaccination and residential mobility.

**Conclusion:** Our results show that different facets of social capital are significantly associated with adoption of protective behaviors, e.g., social distancing, face masking, and vaccination. As such, our results suggest that differential facets of social capital imply a Swiss cheese model of pandemic control planning where, e.g., institutional health and community health, provide partially overlapping behavioral benefits.

## Introduction

Social capital has been developed as a concept to characterize the value of a community structure (1,2). To better reflect the nature of communities, social capital has further been defined as the quality of the relationship among community members, which is represented in trust and reciprocal aid that derive mutual benefits to all parties (3). It has now become widely used to understand social determinants of public health (4,5). In particular, social capital in communities has been associated with health outcomes, such as mortality rate, obesity, and diabetes (6,7) and can explain the variation in health status across different geographic areas (3). For the United States, social capital has been operationalized and measured on county and state levels (8,9).

Social capital is measured using several social elements that each reflect a different aspect of a community (8,9). These facets of social life, such as family unity and institutional trust, can further explain specific social outcomes or behaviors. For example, social capital stemming from family support has been associated with better mental health (10), better mechanisms for coping with stress (11), and lower suicide rates (12). Social capital stemming from civic participation, such as taking part in religious or volunteer groups, promoted better sense of responsibility, and in turn created healthier neighborhoods and higher levels of life satisfaction (13).

### Social Capital and COVID-19

After the declaration of COVID-19 as a pandemic (14), social distancing and wearing masks were recommended as non-pharmaceutical interventions to contain the spread. Even though recommendations were widely announced and justified, not all communities abided uniformly to the new recommendations. Some communities increased an individual sense of responsibility to take actions, e.g., social distancing, to protect self and others (15), whereas other communities found it difficult to isolate and eliminate social gatherings (16,17). Thus, growth of COVID-19 differed among communities and has been associated with social capital and some of its dimensions. The number of COVID-19 confirmed cases decreased with better community health (18), whereases mortality rate increased with lower social capital levels (19), and lack of institutional trust and civic engagement (17). Collective adherence to protective behaviors during a pandemic might mitigate the critical consequences of its spread.

### Physical Distancing

Since COVID-19 is highly infectious and transmits easily with face-to-face interaction, social distancing proved to be an effective mitigation strategy to contain COVID-19 spread (20). Cases decrease by 48% and fatalities by 60% three weeks after states implemented lockdown orders (21). Physical distancing strategies took a variety of forms, from limiting people’s gatherings to fully restricting movements by lockdown orders. In the United States, there were distinctive patterns in mobility reduction among different sociodemographic groups, where some communities voluntarily stayed at home and limited their movements even more after lockdown orders (22).

### Wearing Masks

Face masking also has been an effective non-pharmaceutical intervention which lowers the risks of testing positive for COVID-19 infection by 70% (23,24). In April 2020, with the absence of vaccines, the Centers for Disease Control and Prevention (CDC) recommended the use of cloth masks in public (25), especially after finding that infectious microbes of COVID-19 can be transmitted from persons without symptoms (26,27). All sociodemographic groups adopted mask wearing but there were larger increases in specific geographic areas such as the Midwest, and 76% of the population wears masks when leaving their homes (28).

### Vaccination

For pandemic extinction, it is believed that 70% to 80% of the population must be vaccinated (29,30) and the threshold decreases with following protective health behaviors, such as face masking and social distancing (31). At the initial stage of vaccination development in 2020, half of the U.S. population did not intend to take the vaccine because of health concerns and low confidence in the vaccine (32,33). Also, media misinformation about vaccination strongly lowered people’s intention to be vaccinated, and some sociodemographic groups were impacted differently (34). However, the hesitancy against vaccination started to decline by May 2021 among all demographic groups (35). As hesitancy declined, about 51% of the U.S. population are fully vaccinated as of August 2021(36).

Social responses toward pandemics are critical in containing the spread and mitigating its exacerbated effects. The way communities are structured impacts individuals’ ability to adopt new behaviors, and hence, follow public health recommendations (37). This study aims to explore the association, if any, between different social capital facets and public health behaviors: social distancing, wearing masks and vaccination, during the COVID-19 pandemic at the county-level in the United States.

## Data and Methods

We estimate the effects of social capital on public health behaviors related to COVID-19 pandemic. Table 1 summarizes the variables we used in the analysis with descriptive statistics at the county-level.

**Table 1:**
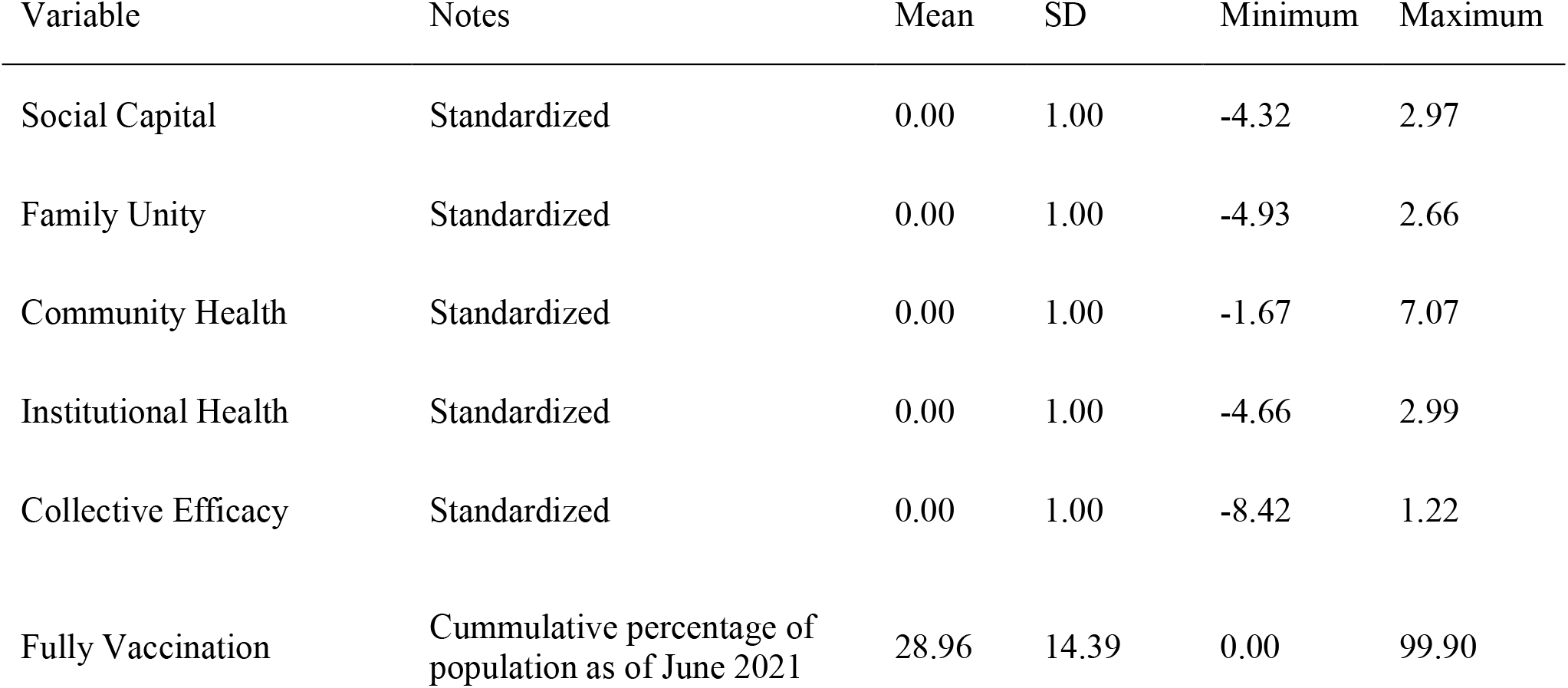
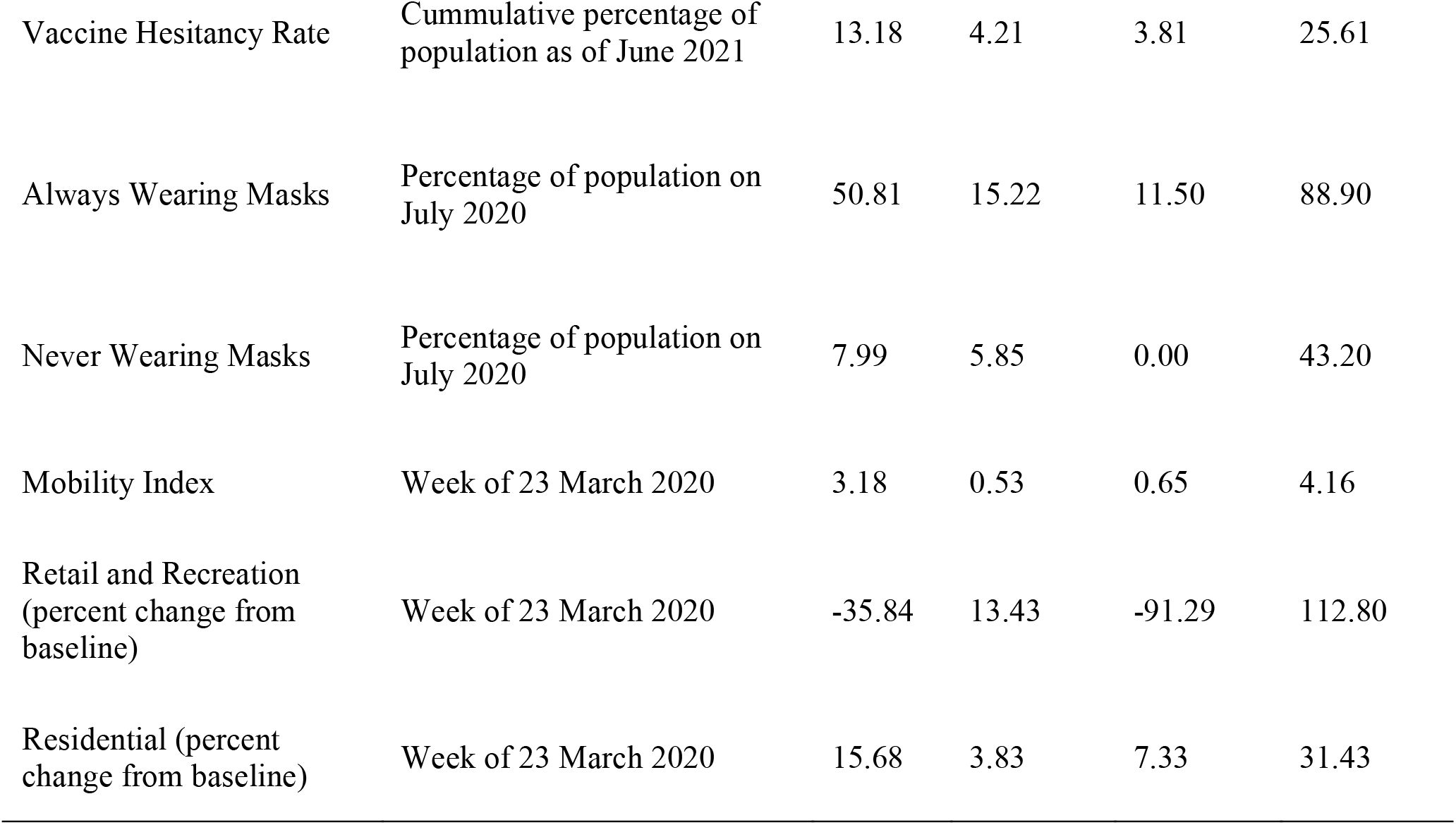
List of variables and descriptives

We obtained county-level social capital indices from the Social Capital Project (9), which comprise four subindices: *family unity* which considers the structure of families in terms of marriage and children; *community health* which considers participation in civic life such as involvement in volunteering and religious groups; *institutional health* which considers confidence in media/corporations/schools, and participation in institutions such as elections and census; and *collective efficacy* which is the converse of social disorganization, operationalized via violent crime rates.

To assess public health behavior, we considered vaccination rate, vaccination hesitancy, mask usage, and changes in mobility patterns at relevant times during COVID-19. We used county-level data considering the fully vaccinated population (38), and estimated vaccine hesitancy (36) as of June 2021. County-level mask usage data is based on a survey of 250,000 people conducted between July 2-14, 2020 (39): we consider extreme responses of “never” and “always”. County-level mobility index is computed by Cuebiq firm (40) based on changes in mobile phone movement. Changes in retail and recreation, and residential mobility are obtained from Google Community Mobility Reports (41). We consider the March 23, 2020 week for mobility data before wide mandates of lockdowns were issued.

The bivariate relationships between each social capital index and public health behavior are assessed using standardized linear regression. Statistical analysis along with P-values and 95% confidence intervals are reported. All analyses have been done using statsmodels package in Python (42).

Datasets used in this study are publicly available (except Cuebiq data) and using them in this research does comply with their owners’ terms and conditions. Cuebiq data is restricted and to request its access one can apply here: https://www.cuebiq.com/.

## Results

Social capital subindices associate differently with COVID-19 vaccination, masking, and mobility change behaviors, illustrated in the Swiss cheese model (43,44) that we extend to social capital (Fig 1). Each public behavior affected by different social capital facets resembles a defensive layer against the spread of COVID-19. The Swiss cheese model is created from the radar charts shown in Fig 2.

**Figure 1:**
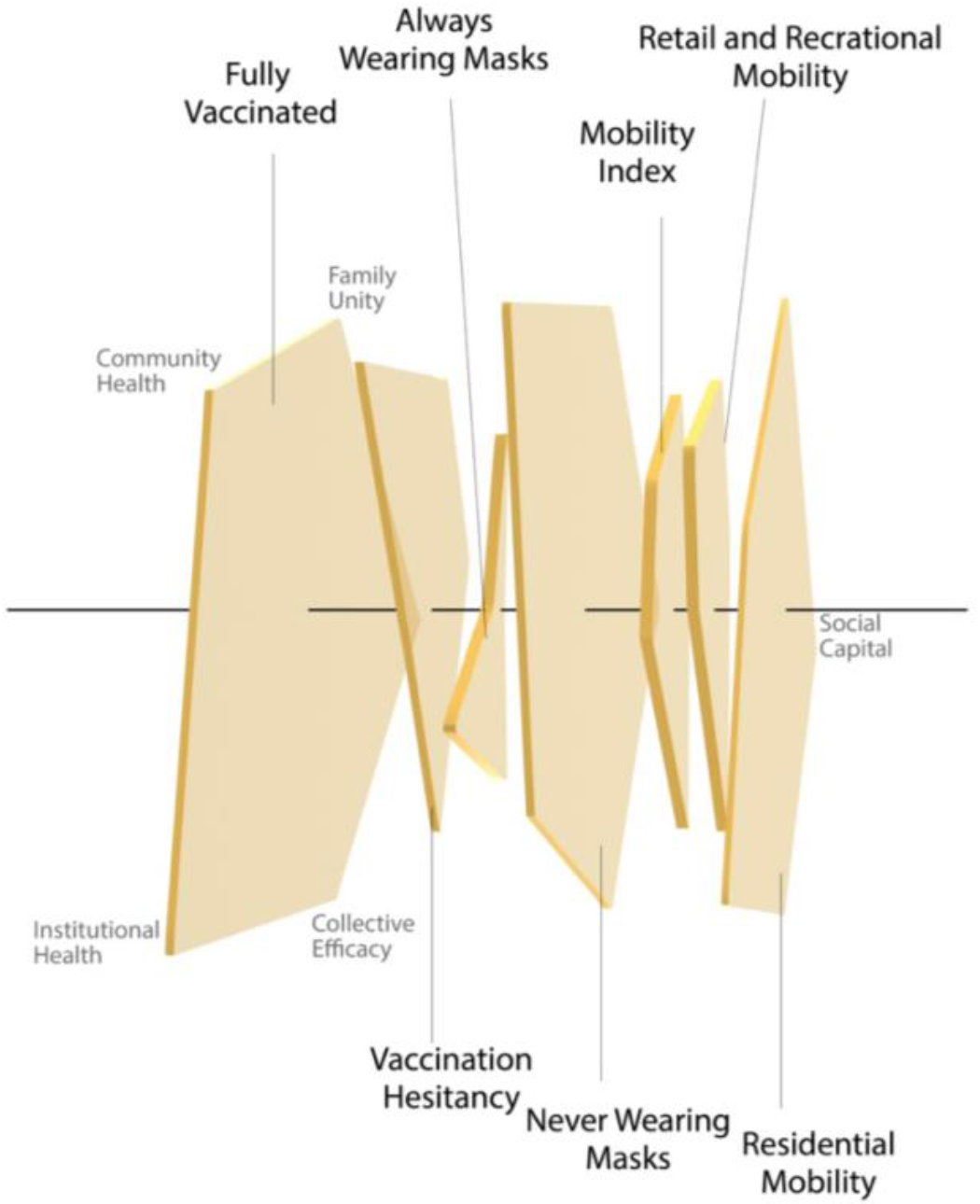
A Swiss cheese model for social capital, with pentagons representing the five social capital indices’ impact on social behaviors

**Figure 2:**
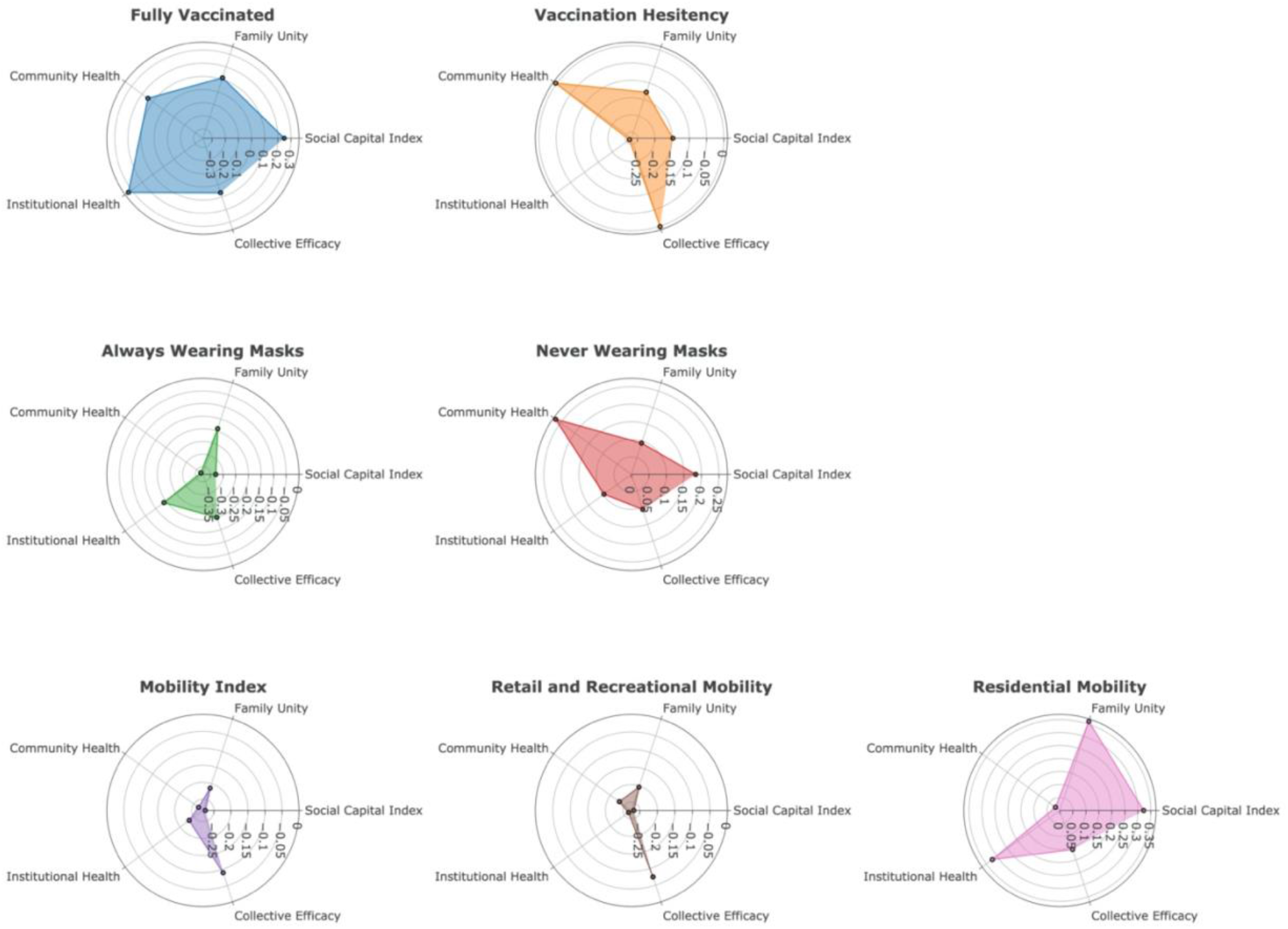
Details of bivariate correlation for each public behavior against social capital indices represented in radar charts

Fig 2 specifies the differences in correlation sizes for each public health behavior against social capital and its four subindices. Correlation coefficients of full vaccination are the largest, and of residential mobility are the second largest, whereas mobility and mask usage have smaller effect sizes. Family unity has similar effect sizes for all public health behaviors except largest for the mobility index. Community health has greatest effect size for masking, less for vaccination, and least for the mobility indices. Institutional health has greatest effect size for vaccination and change in residential mobility but smallest in masking. Collective efficacy has smallest effect size for mobility and largest effect size for mask wearing.

Vaccination mostly associates significantly with institutional health, positively with fully vaccinated population, but negatively with increased hesitant population (Table 2). Fig 3-A shows that over time, counties with high institutional health have an increasing rate of vaccination unlike counties with lower institutional health. Further, Fig 3-B shows that hesitant population is less in counties with higher institutional health.

**Table 2:**
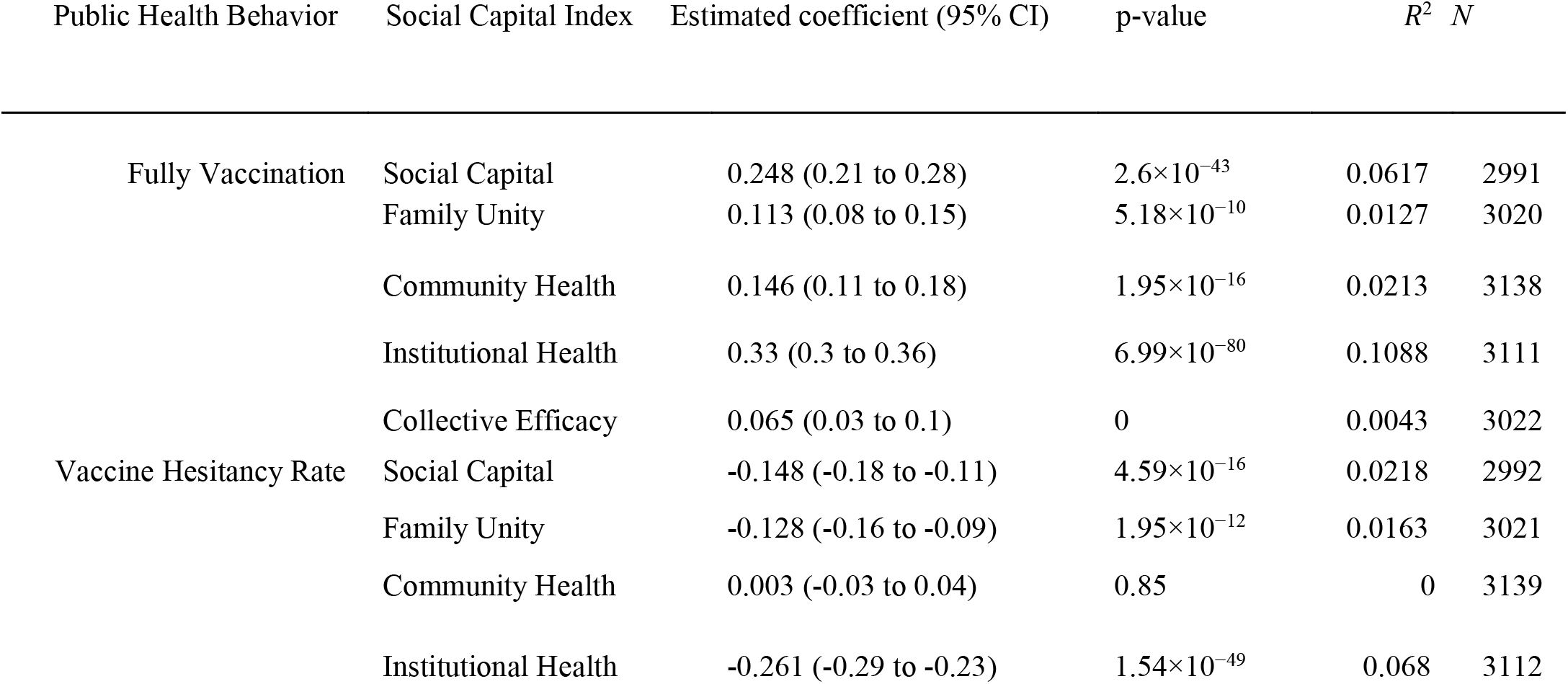
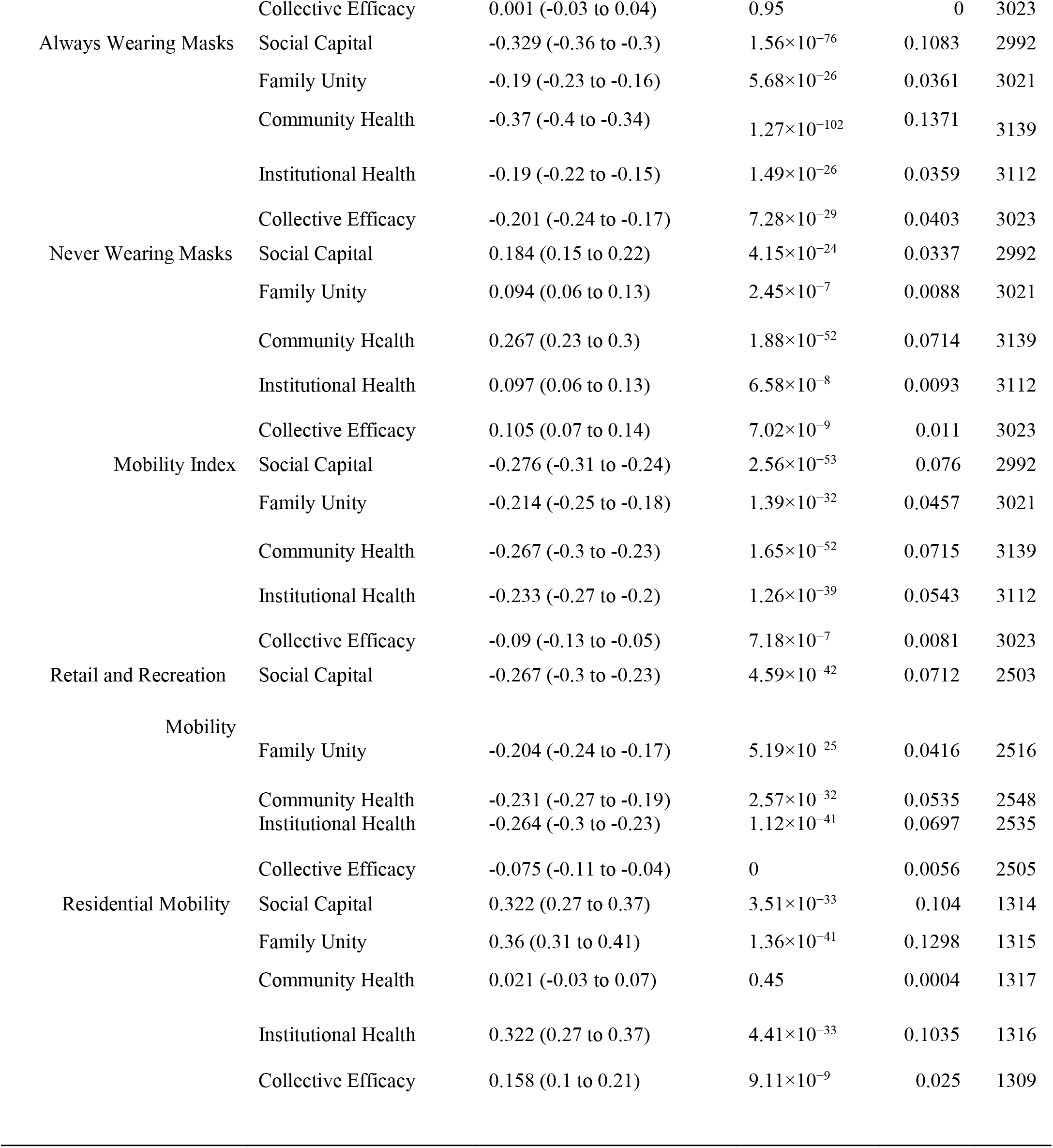
Bivariate correlation estimation table. Each line represents a regression analysis between a public health behavior and social capital index

**Figure 3:**
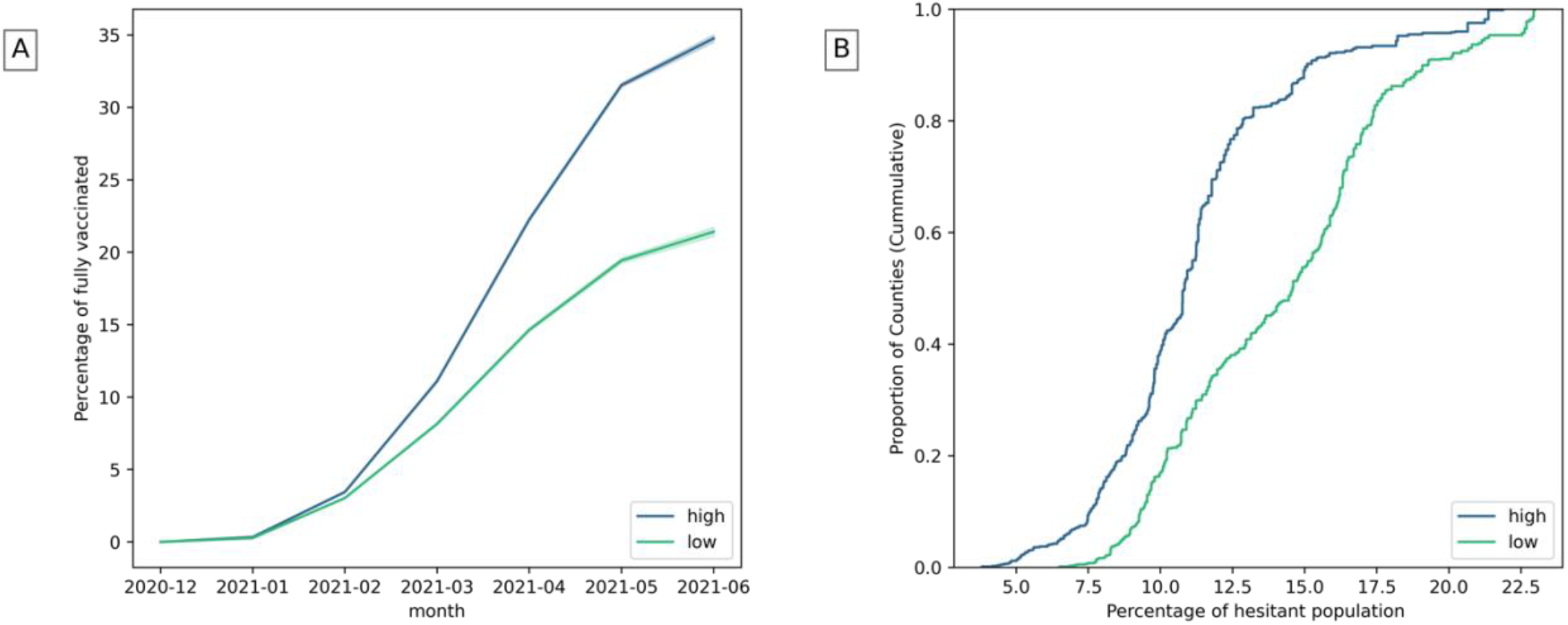
A) Fully vaccinated population over time in counties with high and low institutional health. B) Estimated hesitant population in counties with high and low institutional health. (The highest and lowest 25% of counties are considered).

Most counties have individuals who constantly wear masks, while fewer counties have people who rarely wear masks (Fig 4-A). Wearing masks mostly associate with community health, positively with reduced masks usage and negatively with widely mask usage (Table 2). Fig 4-B shows that counties with higher community health have less people who always wear masks.

**Figure 4:**
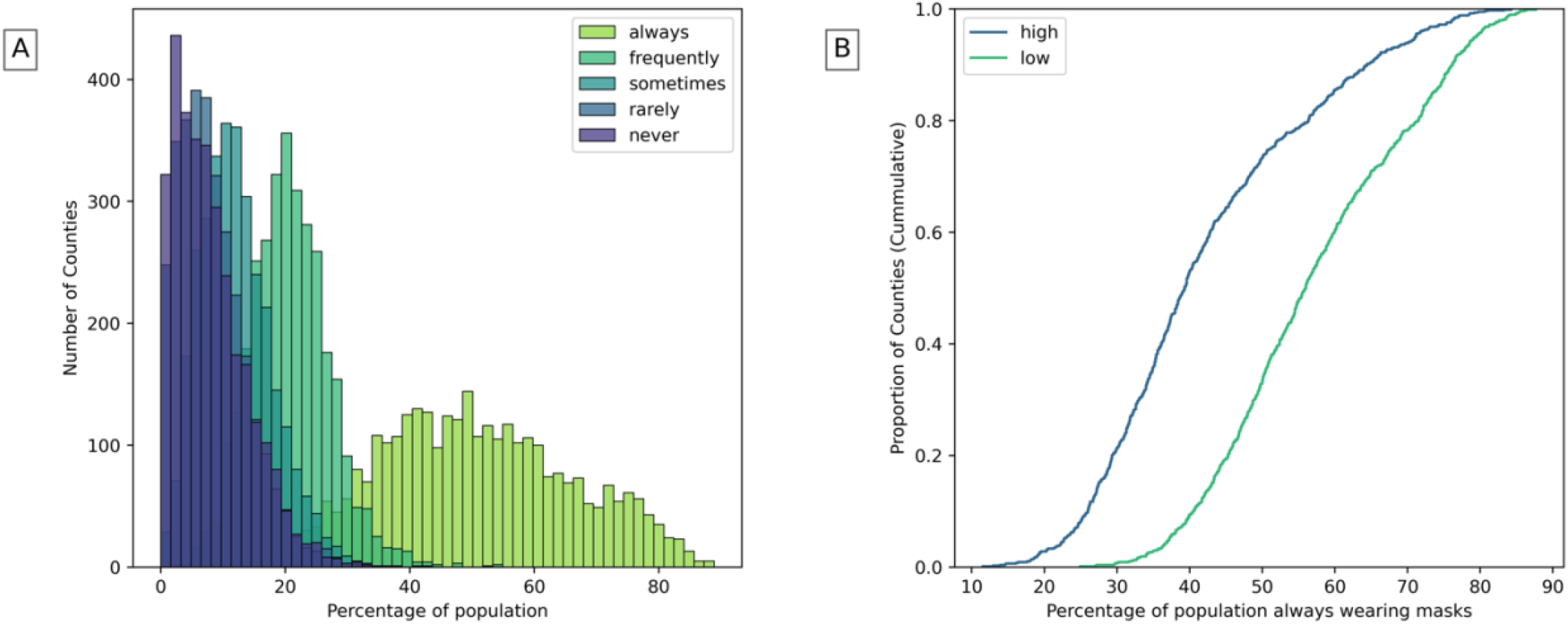
A) Population that wears masks in counties. B) Population that always wears masks in counties with high and low community health (The highest and lowest 25% of counties are considered).

In general, people reduced their visits in recreational areas more than in residential areas (Fig 5-A). Reduced mobility associates mostly with higher overall social capital and secondly with community health (Table 2). Fig 5-B shows that counties with better community health tend to move less. Reduced recreational mobility, as well, associate mostly with higher overall social capital (Table 2) and secondly with better institutional health (Fig 5-C). While increased residential mobility associates mostly with higher family unity (Table 2) as seen in Fig 5-D.

**Figure 5:**
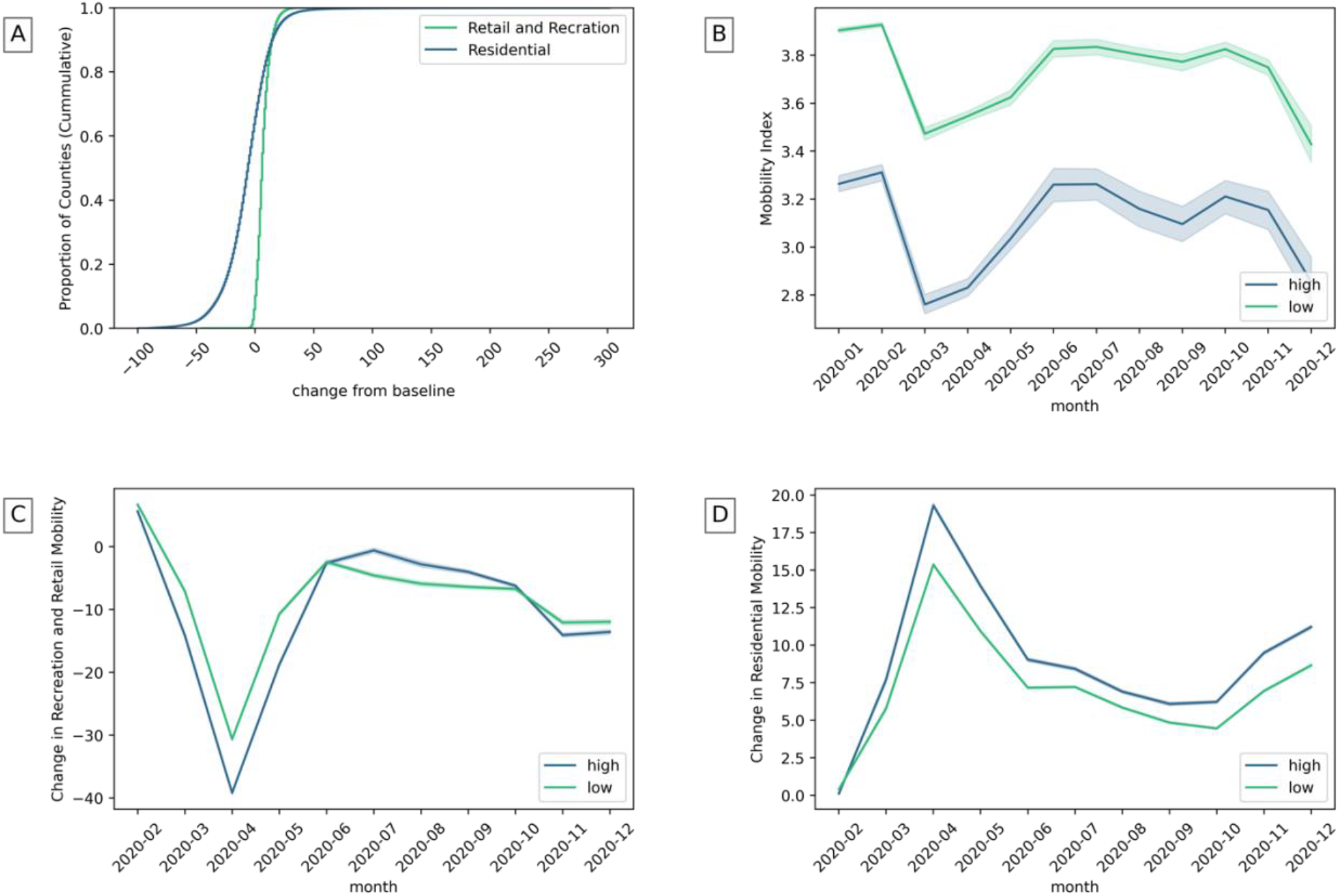
Fig 5: A) The change of recreation and residential mobility in counties. B) Mobility index in counties with high and low community health. C) Recreation and retail mobility in counties with high and low institutional health. D) Residential mobility with high and low family unity. (The highest and lowest 25% of counties are considered).

## Discussion

During pandemics, social capital and its dimensions play a role in differentiating public responses towards health policies and interventions, and in turn health outcomes diverge. Trusting institutions reduced anxiety during SARS pandemic (45), and predicted vaccination acceptance during H1N1 pandemic (46,47). Also, people’s intentions to wearing masks and washing hands increased with better social capital (7).

Similarly, during the COVID-19 pandemic, behavioral responses such as vaccination, masking, and physical distancing have differed among United States counties. Here we have shown that facets of social capital are associated with behavioral responses to the COVID-19 pandemic in different ways. Our findings show that trusting institutions may promote vaccination and reduce vaccination hesitancy. Also, communities with more engagement in civic life tend to reduce their mobility. This supports the findings of Bargain and Aminjonov (2020), Barrios et al. (2021), and Durante et al. (2021), where communities with higher civic life engagement increased the sense of responsibility in individuals to lower their gatherings. However, higher community health may motivate people to relax their face masking, and this can be explained with overall reduced mobility and less face-to-face interactions. People stay home with better family unity whereas recreational visits decrease with better social capital and institutional health.

Our results suggest that social capital and its subindices are essential in explaining differences in public behaviors during health crises which may help determine policies in local communities. Further, our results show that differential facets of social capital imply a Swiss cheese model of pandemic control planning where multiple layers of public behaviors differently affected by social capital can act against contagion spread. There might be some barriers, such as community structure, misinformation from media, and medical concerns, for an individual to adopt new behaviors in pandemics. Therefore, more effort might be needed to help individuals to adhere to new protective behaviors especially in communities that generally have lower social capital.

## Data Availability

https://data.cdc.gov/Vaccinations/COVID-19-Vaccinations-in-the-United-States-Jurisdi/unsk-b7fc

https://github.com/nytimes/covid-19-data/blob/master/mask-use/README.md

https://www.cuebiq.com/

https://www.google.com/covid19/mobility/

https://data.cdc.gov/Vaccinations/Vaccine-Hesitancy-for-COVID-19-County-and-local-es/q9mh-h2tw

https://www.jec.senate.gov/public/index.cfm/republicans/2018/4/the-geography-of-social-capital-in-america

